# Scenarios for the long-term efficacy of amyloid-targeting therapies in the context of the natural history of Alzheimer’s disease

**DOI:** 10.1101/2024.02.26.24303371

**Authors:** Lars Lau Raket, Jeffrey Cummings, Alexis Moscoso, Nicolas Villain, Michael Schöll

**Affiliations:** Eli Lilly and Company, Indianapolis, Indiana; Clinical Memory Research Unit, Department of Clinical Sciences, Lund University, Lund, Sweden; Chambers-Grundy Center for Transformative Neuroscience, Pam Quirk Brain Health and Biomarker Laboratory, Department of Brain Health, School of Integrated Health Sciences, University of Nevada Las Vegas (UNLV), Las Vegas, NV, USA; Wallenberg Centre for Molecular and Translational Medicine and the Department of Psychiatry and Neurochemistry, University of Gothenburg, Sweden; AP-HP Sorbonne Université, Pitié-Salpêtrière Hospital, Department of Neurology, Institute of Memory and Alzheimer’s Disease, Paris, France; Sorbonne Université, INSERM U1127, CNRS 7225, Institut du Cerveau - ICM, Paris, France; Department of Clinical Physiology, Sahlgrenska University Hospital, Gothenburg Sweden; Dementia Research Centre, Queen Square Institute of Neurology, University College London, United Kingdom

## Abstract

**Importance:** Recent clinical trials of Aβ-targeting therapies in Alzheimer’s disease (AD) have demonstrated clinical benefit over 18-months, but their long-term impact on disease trajectory is not yet understood. We propose a framework for evaluating realistic long-term scenarios.

**Objective:** To integrate data from recent phase 3 trials of the high-clearance Aβ-targeting antibodies with an estimate of the long-term patient-level natural history trajectory of the Clinical Dementia Rating-Sum of Boxes (CDR-SB) score, to explore realistic long-term efficacy scenarios.

**Data Sources:** Published results from recent phase 3 randomized clinical trials of high-clearance Aβ-targeting antibodies with completion between 2019 and 2023.

**Study Selection:** All phase 3 studies of aducanumab, lecanemab, gantenerumab, and donanemab were included.

**Main Outcomes and Measures:** Reported study results on CDR-SB estimated using categorical-time models.

**Results:** Three distinct long-term efficacy scenarios were examined, ranging from conservative (enduring short-term delay), over intermediate (fading stage-dependent slowing), to optimistic (continued stage-independent slowing). In a hypothetical framework, we found that even modestly effective therapies with fading stage-dependent slowing could result in major delays of disease progression if initiated in the earliest stages of AD. In long-term scenarios for the treatment effects observed in recent positive phase 3 trials, we found that initiating the treatments in the early symptomatic stages, defined by the respective trial inclusion criteria, could delay the onset of severe dementia by 0.3-0.6 years (conservative), 1.1-1.9 years (intermediate), and 2.0-4.2 years (optimistic).

**Conclusion and Relevance:** The findings provide insights into hypothetical long-term impact of Aβ-targeting treatments, highlighting the potential of maximizing clinical benefit with earlier intervention. This study underscores some of the complexities of evaluating and comparing Aβ-targeting therapies in the context of AD’s nonlinear disease trajectory and the need for considering both differences in trial populations and duration. Our work calls for studies with longer follow-up and results from early intervention trials to provide a comprehensive assessment of these therapies’ true long-term impact.

## Introduction

Alzheimer’s disease (AD) is a progressive neurodegenerative disease characterized by the pathological accumulation of amyloid-β (Aβ) and tau aggregates in the brain.^1^ Although the exact mechanism of AD pathogenesis remains unclear, current neuropathologic, genetic, and human *in vivo* studies strongly support the “*amyloid cascade hypothesis*”^2^ which posits Aβ as a key agent in the pathologic process of AD. Aβ has thus been among the most common therapeutic targets of experimental drugs for patients with AD.^3^

After years of failed attempts to develop clinically effective Aβ-targeting therapies, converging evidence indicates that therapies targeting Aβ aggregates that reduce brain Aβ plaque load below ∼20-25 centiloids on Aβ positron emission tomography (PET), i.e., the threshold commonly used to define “Aβ-positivity”, has clinical benefits for patients with early symptomatic AD.^4^ This is evidenced by statistically significant reductions of 22-36% in decline in the Clinical Dementia Rating–Sum of Boxes (CDR-SB) score across the EMERGE (aducanumab),^5^ CLARITY AD (lecanemab),^6^ and TRAILBLAZER-ALZ 2 (donanemab)^7^ trials, with associated amyloid clearance rates of 48%, 68% and 80%. The confirmatory trials ENGAGE (aducanumab) and GRADUATE I and II (gantenerumab) did show significant amyloid reductions with amyloid clearance rates of 31%, 28% and 27%,^5,8^ but failed to show statistically significant clinical benefits with observed reductions in decline of −2%, 8% and 6%. However, an exploratory post-hoc analysis in the GRADUATE I and II trials of gantenerumab suggested that these negative outcomes reflected the failure to achieve the desired degree of Aβ plaque removal.^8^

While these results represent a milestone in the search for effective disease-modifying therapies, the clinical and economic relevance of the observed treatment effects, particularly beyond the 18-month trial duration, remain a subject of debate and carry implications for decisions about the potential implementation and reimbursement of these treatments.^9–12^

To better understand the significance of the novel Aβ-targeting therapies, it is crucial to interpret treatment effects in the context of the natural history of AD. Abnormal Aβ accumulation is a slow process that begins years before the onset of symptoms,^13^ as reflected by the fact that many cognitively unimpaired (CU) older individuals (∼30% over 70 years of age) have elevated Aβ plaque loads in their brain (and are thus defined as Aβ-positive).^14^ While this phase of Aβ-positive CU status can last many years, it is associated with a greatly increased risk of progressing to symptomatic Alzheimer’s disease over the long term.^15,16^ Most trials of Aβ-targeting therapies, including the three positive confirmatory trials to date, have primarily focused on Aβ-positive subjects with early symptomatic AD which increases the likelihood of demonstrating a clinical benefit over a shorter time span compared to trials with Aβ-positive cognitively unimpaired individuals. There is a hypothesis that addressing Aβ-pathology before symptoms could delay or entirely prevent cognitive decline due to AD.^17^ This hypothesis has yet to be tested for the Aβ-targeting monoclonal antibodies that have demonstrated efficacy in the early symptomatic stages of AD, but trials with longer-term follow-up in presymptomatic individuals are underway.^18,19^

This paper addresses scenarios for long-term treatment effects of recent Aβ-targeting therapies. To address the question in a framework that is consistent with AD progression, we used an estimate of the typical patient-level natural history trajectory of CDR-SB from the earliest stages of preclinical AD to severe AD dementia. The estimated trajectory was based on data from the Alzheimer’s Disease Neuroimaging Initiative (ADNI) using a previously validated disease progression model.^20^ This long-term reference trajectory allowed for the exploration of different scenarios for long-term treatment effects. We used published data from the recent trials of high-potency Aβ-targeting therapies to model long-term treatment effects relative to the natural history trajectory of CDR-SB. The results of our study contribute to elucidating the potential of Aβ-targeting therapies to produce and maintain clinically meaningful benefits over varying time durations.

## Methods

### Clinical studies and efficacy results

This study included data from the following recent phase 3 trials of high-clearance Aβ-targeting antibodies: EMERGE and ENGAGE of aducanumab, CLARITY AD of lecanemab, GRADUATE I and II of gantenerumab, and TRAILBLAZER-ALZ 2 of donanemab. The trials included in this analysis had slightly different entry criteria. For example, the range of allowable Mini-Mental Status Examination (MMSE) scores at baseline differed among studies. All the trials involved early AD with overlapping population characteristics.

The EMERGE and ENGAGE studies included both a high and a low dose of aducanumab, however, since the high dose is recommended^21^ the low dose was not considered here. Since the ENGAGE study did not yield numerical benefit of high-dose aducanumab before being terminated for futility (0.02 CDR-SB points worsening *vs.* placebo, p = 0.833),^5^ the long-term extrapolations based on this study shows no benefit compared to placebo; hence, these results will not be displayed in figures in the main manuscript. For completeness, figures showing results for EMERGE and a combined analysis across EMERGE and ENGAGE are reported in the Supplementary Material. The primary patient population in TRAILBLAZER-ALZ 2 had a low/medium tau load based on [^18^F]flortaucipir PET retention patterns and quantification (standardized uptake value ratios, SUVR). In the present analysis, we focused on this population. We report comparisons of results of the primary low/medium tau population and the combined low/medium and high tau population in the Supplementary Material.

Baseline scores and trajectories of change in CDR-SB and associated standard deviations were extracted from publications or public presentations. For comparability, we used trajectories estimated using categorical time models. Most studies reported results based on the mixed model for repeated measures (MMRM), but results for the GRADUATE I & II studies were reported based on a reference-based multiple imputation ANCOVA model that is closely related to MMRM. All results were reported as estimated marginal means representing an average patient in the trial. For all studies, the CDR-SB results at the final visits were reported, but some intermediate data points required extraction from graphs using WebPlotDigitizer.^22^

The time delay at the final visit and associated time saving (time delay divided by trial duration) were estimated from the extracted MMRM trajectories by using natural cubic spline interpolation of the placebo results and computing the time at which the placebo trajectory crossed the score of the active arm at the final visit.

### Natural history trajectory of CDR-SB

A long-term patient-level natural history trajectory of CDR-SB from the earliest stages of preclinical AD to late-stage dementia was estimated based on a previously published disease progression model.^20^ The detailed specification of the model and estimated trajectory has been described.^23^ Briefly, the model is a latent-time disease progression model that simultaneously estimated trajectories of CDR-SB, ADAS-cog (13-item), MMSE and Aβ PET (centiloids) based on longitudinal observations (4581 follow-up years) from 1424 well-characterized subjects from ADNI. Subjects were either Aβ negative and cognitively unimpaired or Aβ positive with any cognitive status. Based on the continuous-time staging of patients, the natural history CDR-SB trajectory was estimated based on all available observations of CDR-SB using a mixed-effects model with the mean trajectory modeled as a natural cubic spline model with 5 degrees of freedom and a patient-level random intercept. The estimated CDR-SB trajectory used for this study is specified in Table S1 in the Supplementary Material.

We explored the effects of intervening at different disease stages with hypothetical interventions that resulted in cumulative stage-dependent (fading) and stage-independent slowing of disease progression. Based on placebo trajectories, we predicted the time to severe dementia defined as a CDR-SB score of 16, and the predicted treatment-associated time delays compared to placebo in progressing to severe dementia.

### Integrating clinical trial trajectories with natural history trajectories

We used average baseline CDR-SB scores in the clinical studies to identify the starting points of the trial trajectories along the natural history trajectory of CDR-SB.

The active-arm and placebo-arm trajectories were extrapolated throughout the disease course. For the placebo arms, a post-trial decline proportional to the natural history trajectory was assumed (fixed time shift of natural history trajectory to match the placebo arm at final visit), while for the active arms, three different assumptions were explored:

1. **Enduring short-term delay.** The time delay of progression relative to placebo observed at the end of the double-blind treatment exposure is enduring, but after that, disease continues to follow the natural history trajectory. For example, if 6 months delay in disease progression was observed at the end of the trial, all subsequent milestones occur with a 6 month delay relative to the extrapolated placebo trajectory.
2. **Stage-dependent fading slowing.** The time saving observed at the end of the trial (time delay divided by trial duration) was assumed to continue at a decaying rate past the end of the trial. The rate of time saving was assumed to decrease linearly from the observed slowing at end of the trial to 0% time saving once the trajectory reached CDR-SB = 16, at which point the trajectory continued to follow the natural history trajectory. For example, if a 6 months delay in disease progression was observed at the end of an 18-month trial (33%), and it took 72 months from baseline for the extrapolated active treatment arm trajectory to reach CDR-SB = 16, the delays at 36, 54, and 72 months after treatment initiation would be 11 months (31%), 14 months (26%), and 15 months (21%), respectively. After 72 months, no additional increase in the delay would occur.
3. **Stage-independent continued slowing.** The time saving observed at the end of the trial (time delay divided by trial duration) was assumed to continue past the end of the trial, where patients followed the natural history trajectory at a slower pace. For example, if 6 months delay in disease progression was observed at the end of an 18-month trial (33%), disease progression extrapolated by the natural history trajectory would be delayed by 12 months 36 months after treatment initiation (33%).

The first scenario was considered the most conservative disease-modifying scenario, where no additional benefit was accrued past the end of the trial period, while the third scenario was considered optimistic. The second scenario is an intermediate scenario between 1) and 2).

## Results

The adjusted mean CDR-SB trajectories for the five phase 3 studies are shown in Figure 1.

**Figure 1.**
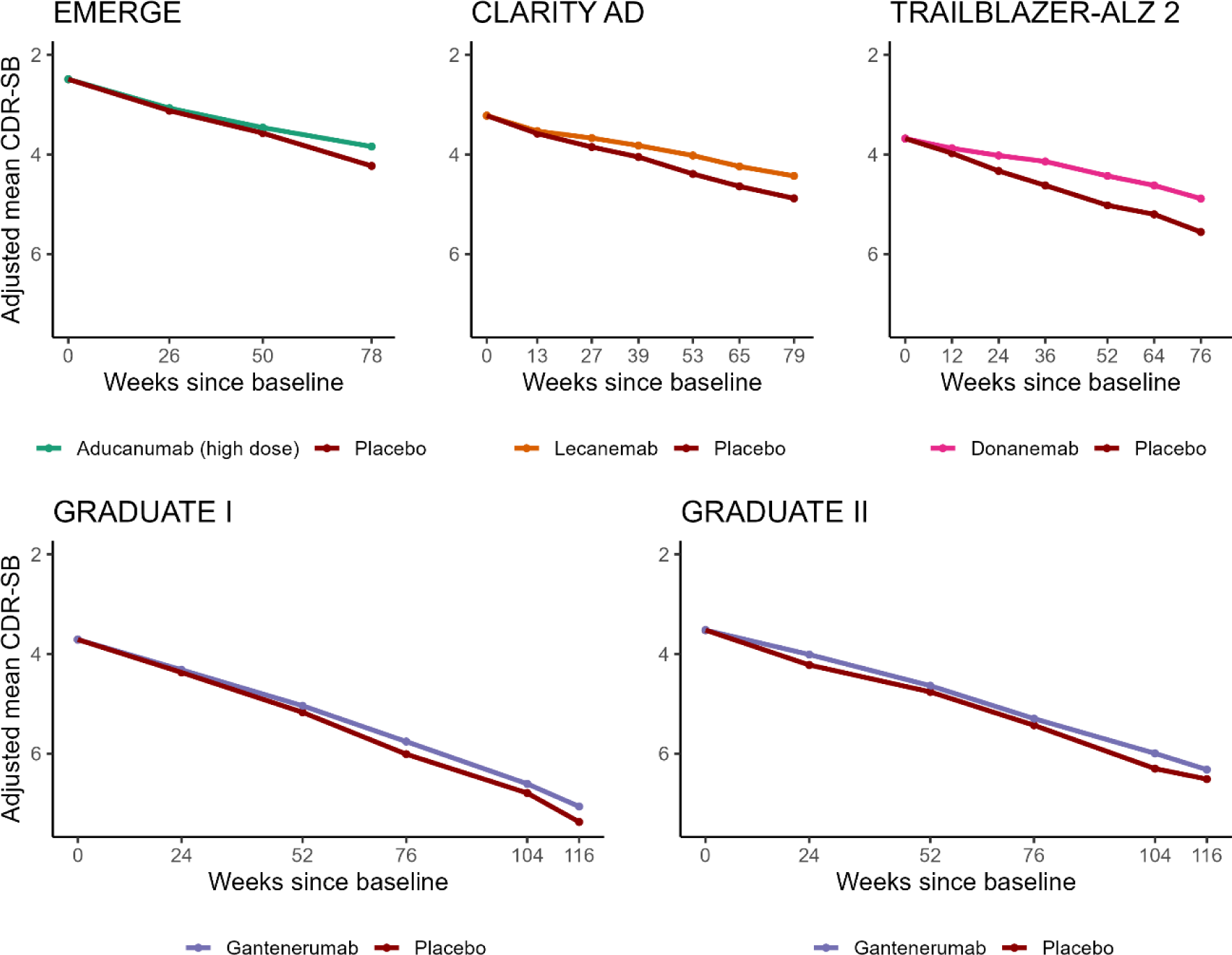
Clinical Dementia Rating sum of boxes (CDR-SB) results of trials and treatment groups included in the present study. Results are based on publicly reported results of change from baseline in CDR-SB analyzed using the mixed model for repeated measures with the reported average baseline score added to the results to bring them to the CDR-SB scale.

The characteristics of the trials, disease severity of the trial populations at baseline, and quantifications of treatment effects (treatment differences, time delays) are shown in Table 1. Disease severity of the trial populations differed in terms of their mean baseline CDR-SB scores and predicted years since Aβ PET positivity of the average participants. The results suggest that the population in EMERGE was the least progressed, followed by CLARITY AD, and then TRAILBLAZER-ALZ 2 and GRADUATE I & II populations.

**Table 1.** Characteristics of trials and estimated treatment effects on CDR-SB at final visit.

| **Trial** | **Treatment** | **Trial duration** | **Mean baseline CDR-SB** | **Predicted time since Aβ PET positivity at baseline** | **CDR-SB difference at final visit** | **Estimated time delay at final visit** | **Estimated time saving at final visit** |
| --- | --- | --- | --- | --- | --- | --- | --- |
| **EMERGE** | Aducanumab | 78 weeks | 2.49 | 10.0 years | -0.39 | 15.7 weeks | 20% |
| **ENGAGE** | Aducanumab | 78 weeks | 2.40 | 9.9 years | 0.03 | -1.1 weeks | -1% |
| **CLARITY AD** | Lecanemab | 79 weeks | 3.22 | 10.7 years | -0.45 | 24.3 weeks | 31% |
| **GRADUATE I** | Gantenerumab | 116 weeks | 3.71 | 11.1 years | -0.31 | 6.1 weeks | 5% |
| **GRADUATE II** | Gantenerumab | 116 weeks | 3.52 | 11.0 years | -0.19 | 11.1 weeks | 10% |
| **TRAILBLAZER-ALZ 2** | Donanemab | 76 weeks | 3.68 | 11.1 years | -0.67 | 30.1 weeks | 40% |

The estimated natural history trajectory of CDR-SB is shown in Figure 2 along with different treatment effect scenarios. Figure 2a illustrates the effect of intervening at different time points with a treatment that results in 30% continued stage-independent slowing and Figure 2b illustrates the effect of intervening at the same time points with a treatment that results in 30% fading stage-dependent slowing. This illustrates the long-term benefit of intervening earlier in disease if the treatment effect accumulates over time. Severe dementia, defined as a CDR-SB score of 16 or greater, occurred 17.7 years after Aβ PET positivity. In the 30% continued slowing scenario, when intervening at the time of becoming Aβ PET positive, severe dementia was delayed by 7.6 years. When intervening 5, 10 and 15 years after amyloid positivity, severe dementia was delayed by 5.4, 3.3 and 1.2 years, respectively. In the 30% fading slowing scenario, the corresponding delays were 3.4, 2.5, 1.6 and 0.7 years.

**Figure 2.**
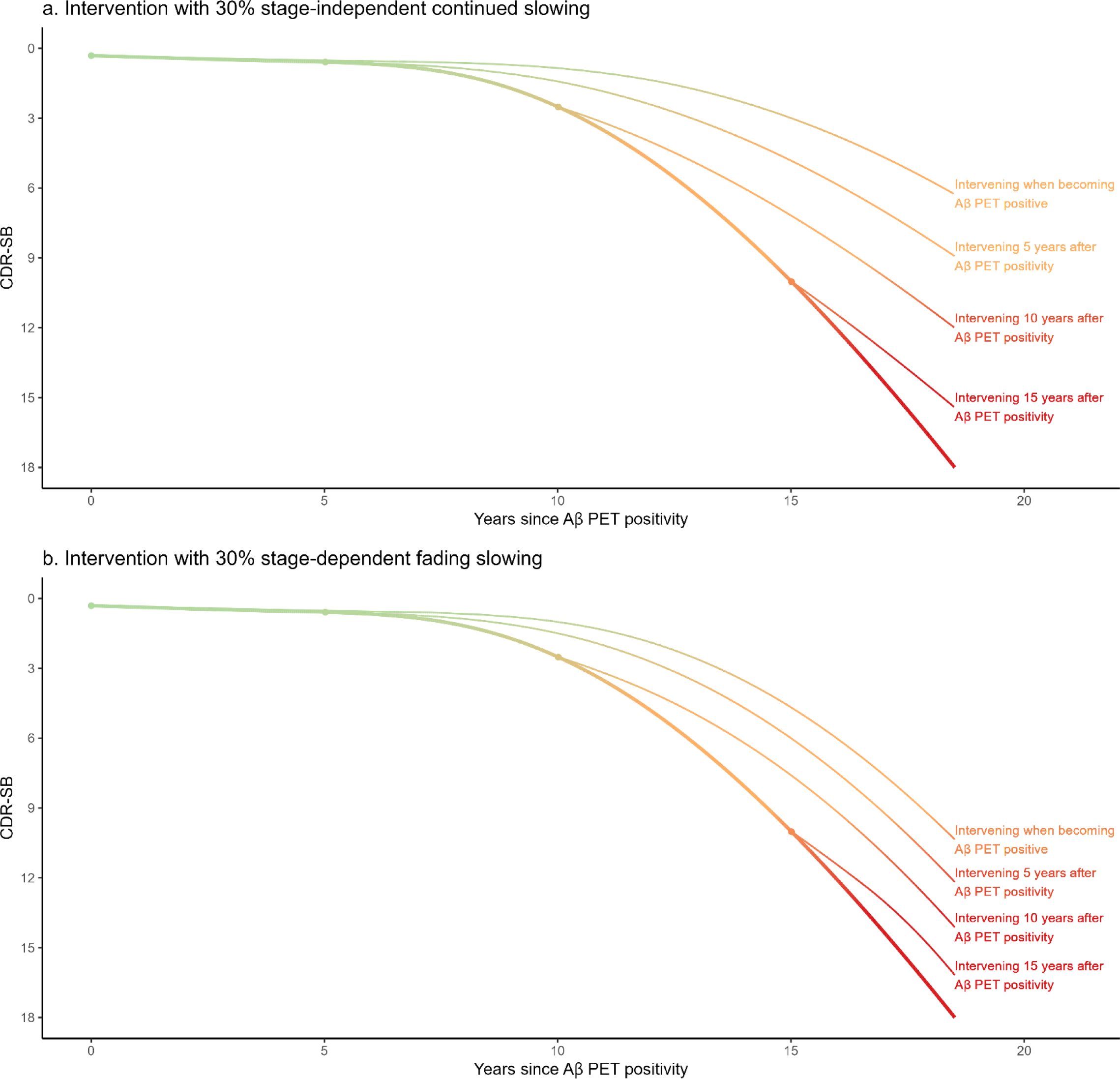
Estimated natural history trajectory of CDR-SB (thick line) from Raket et al.^23^ Four examples (thin lines) illustrate how hypothetical interventions that produces: a) 30% stage-independent continued slowing of disease progression, and b) 30% stage-dependent fading slowing; would change the trajectory based on when the intervention was started (0, 5, 10 or 15 years after Aβ PET positivity).

For each trial, the three long-term efficacy scenarios are shown in Figure 3. Overall, the positive trials (EMERGE, CLARITY AD and TRAILBLAZER-ALZ 2) showed similar long-term trajectories. Despite of the differences in baseline disease stage across trials, the benefit associated with long-term trajectories were consistent with trial results, with TRAILBLAZER-ALZ 2 (most progressed patients) showing the best long-term results, followed by CLARITY AD and EMERGE (least progressed). Consistent with the results, the GRADUATE I and II trials and ENGAGE (Figure S2) showed the least long-term benefit in all scenarios.

**Figure 3.**
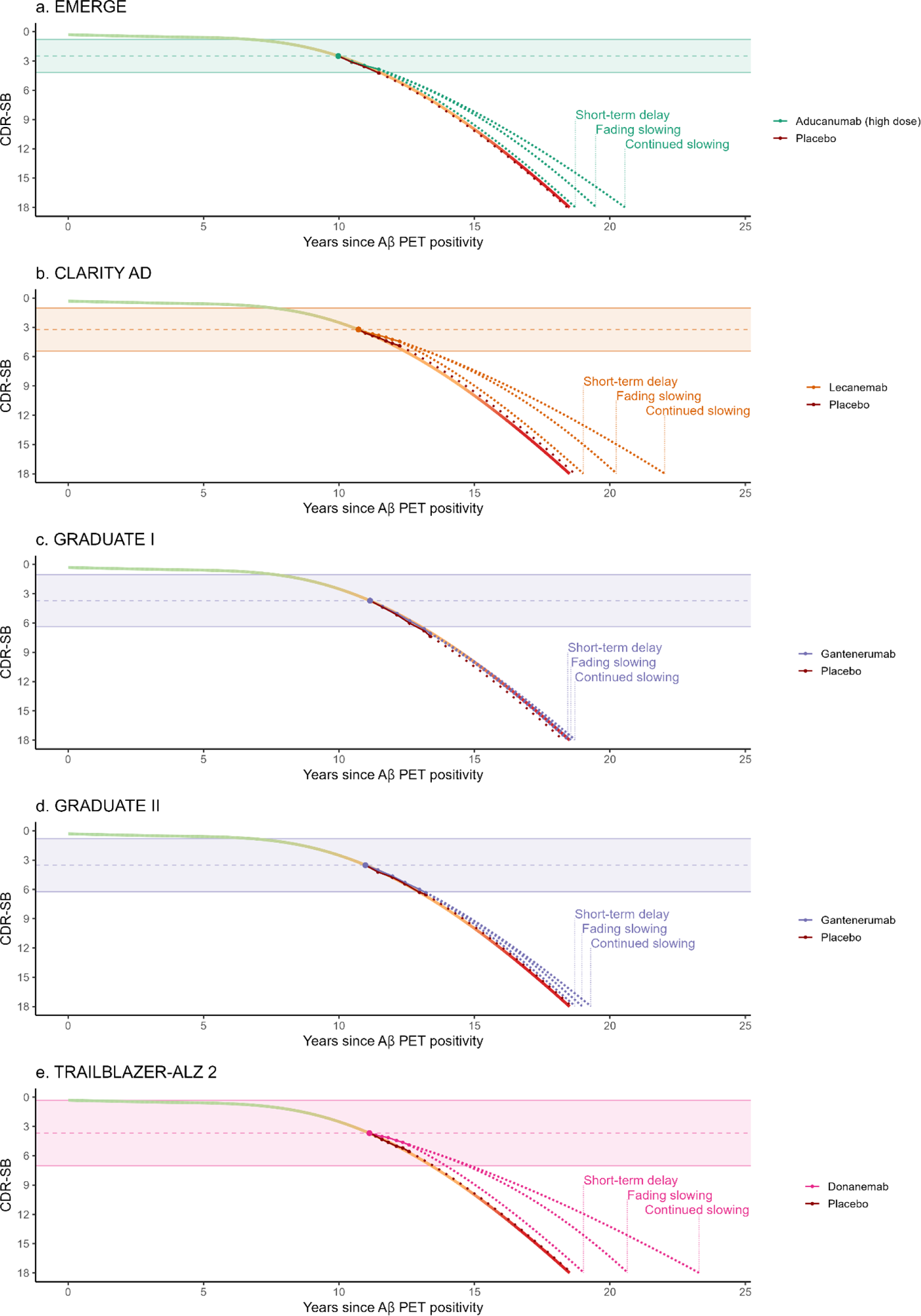
Trial results mapped to the long-term CDR-SB trajectory with extrapolations (dotted lines). The gradient trajectory represents the estimated natural history trajectory, and the shaded bands represent the 90% prediction intervals for baseline CDR-SB scores in the different studies.

The projected CDR-SB differences to placebo and associated time delays of disease progression at 3, 4 and, 5 years after treatment initiation in the individual trials are given in Table S3 in the Supplementary Material. The estimated delays in time to severe dementia are given in Table 2.

**Table 2.** Extrapolated estimates of time to severe dementia (CDR-SB = 16).

| Trial | Treatment | Time to severe dementia (placebo) | Treatment-associated delay in time to severe dementia |  |  |
| --- | --- | --- | --- | --- | --- |
|  |  |  | Short-term delay | Fading slowing | Continued slowing |
| **EMERGE** | Aducanumab | 7.6 years | 0.3 years | 1.1 years | 2.0 years |
| **ENGAGE** | Aducanumab | 7.9 years | −0.0 years | −0.1 years | −0.2 years |
| **CLARITY AD** | Lecanemab | 7.2 years | 0.5 years | 1.4 years | 2.9 years |
| **GRADUATE I** | Gantenerumab | 6.3 years | 0.1 years | 0.5 years | 0.6 years |
| **GRADUATE II** | Gantenerumab | 6.8 years | 0.2 years | 0.4 years | 0.6 years |
| **TRAILBLAZER-ALZ 2** | Donanemab | 6.6 years | 0.6 years | 1.9 years | 4.2 years |

Comparisons of EMERGE and ENGAGE and a combined analysis are shown in Figure S1 and S2 in the Supplementary Material. Comparisons of the primary low/medium tau population and the combined low/medium/high tau population in TRAILBLAZER-ALZ 2 are given in Table S2 and Figures S3 and S4 in the Supplementary Material.

## Discussion

This study explored different long-term efficacy scenarios of Aβ-targeting therapies in the context of the natural history trajectory of AD. The findings shed light on some of the challenges and complexities of evaluating and comparing the clinical efficacy of Aβ-targeting therapies based on the currently available trial results. The modeling presented here highlights the importance of comparing trial results in a manner that considers both differences in trial populations and trial duration, which will influence the observed treatment differences due to the highly nonlinear natural history trajectory of CDR-SB over the course of AD.

Among our three long-term scenarios, one was considered conservative and one optimistic. The conservative scenario assumed that the benefit accrued during the trial period was lasting, but that the disease would progress according to the natural history trajectory with no additional benefit after the trial period. The optimistic scenario extrapolated the time saving observed during the study period to the remaining disease course. In the optimistic scenario, patients kept declining, but the reason for considering this optimistic is that we found it unlikely that amyloid clearance could result in substantially increasing benefit after the end of the trial periods in symptomatic AD patients, given the existence of pathological tau and co-pathologies such as vascular disease, Lewy bodies and TDP-43 aggregates in the vast majority of these patients.^24^ Based on the currently available data, it is our hypothesis that amyloid-clearance in the early symptomatic stages of AD is likely to result in long-term trajectories within the spans of the conservative and optimistic long-term trajectories shown in Figure 3. Our scenario with fading stage-dependent slowing is one example of an intermediate scenario, but one can construct infinitely many different scenarios within this span.

The rationale for a continued slowing of disease progression beyond the clinical trial duration is primarily rooted in the observation that amyloid pathology facilitates the spreading of tau pathology,^25^ and thus acts as an accelerant of clinical disease progression. However, due to the “prion-like” nature of pathological tau,^26^ the time saving related to Aβ-targeting therapies could diminish in later stage disease where more widespread tau pathology may be less reliant on amyloid-related pathways to accelerate spreading and propagation.^25,27,28^ Similarly, cell death and accrual of non-Aβ, non-tau pathologies may reflect the shape of the disease trajectory in later years. These observations motivated our scenario with a stage-dependent fading slowing. An assumption that may be challenged is that the slowing observed in the individual trial was an inherent result of the given therapy in the chosen trial population. While the different Aβ-targeting monoclonal antibodies differ in epitopes^29^ which may result in unique properties,^30^ an alternative assumption could be that the observed efficacy is fully driven by the speed and depth of amyloid clearance. This assumption would suggest that the greater time saving seen in the TRAILBLAZER-ALZ 2 trial was driven by the observed faster and deeper amyloid clearance rather than donanemab’s specific target. The implication would be that once a subject reaches a sufficiently low global amyloid load, they will experience the full benefit of amyloid clearance that will be independent of the therapy used.

This study focused on time delays and time saving to avoid some of the pitfalls of working with proportional reduction in decline, which can be unstable when trajectories are nonlinear and lead to unnatural extrapolations (e.g., that patients on active treatment can never fully decline).^31^ However, the focus on time-based treatment effects required us to compute treatment effects based on summary data. Time saving estimates from CLARITY AD based on a mixed-effects model assuming a linear trajectory of patient-level data have been reported previously.^32^ This patient-level analysis estimated that lecanemab delayed progression by 5.3 months after 18 months treatment. This estimate corresponds to 29% time saving, which is very similar to the 31% extracted from the MMRM in the present analysis. In TRAILBLAZER-ALZ 2, it was reported that donanemab resulted in 33 weeks delay of disease progression after 76 weeks treatment corresponding to a 43% time saving.^7^ These results were based on a proportional time saving Progression Model for Repeated Measures (PMRM) analysis of patient-level data.^31^ The time delay estimate based on the MMRM trajectories that was used for the present analysis was similar with an estimated 40% time saving.

In addition to the limitations of the long-term extrapolation scenarios mentioned previously, the study also has limitations in terms of the anchoring of studies on the estimated CDR-SB natural history trajectory. The different studies had different inclusion criteria, which may lead to biased sampling of subjects along the disease timeline, causing the trial trajectories not to align with the estimated natural progression. In the studies analyzed, we found that the placebo groups’ progression patterns closely matched the estimated natural history, with the greatest deviation in the combined low/medium/high tau population in TRAILBLAZER-ALZ 2 (Figure S4). Furthermore, the analyses in the present study were based on matching adjusted mean trajectories from trials to a trajectory that was estimated to represent a typical patient with AD. Caution is warranted when interpreting these results in relation to individual patient-level trajectories, where factors such as AD pathological load, co-pathologies and cognitive reserve may influence rate of decline and potentially the response to treatment.^33–35^

In summary, the Aβ-targeting treatments that have shown significant impact on clinical outcomes over 18 months delayed disease progression on CDR-SB between 4 and 7 months during the study periods. Based on the long-term scenarios examined in this study, this may lead to delays of severe dementia by 4-7 months (conservative), 1.1-1.9 years (intermediate), or 2.0-4.2 years (optimistic) if treatment is initiated in the early symptomatic stages of AD. If amyloid clearance results in cumulative benefits like observed in the continued and fading slowing scenarios, the most substantial long-term benefits would occur if using Aβ-targeting treatments in the presymptomatic stage of AD.

## Data Availability

All data produced in the present work are contained in the manuscript

## Supplementary Material

**Table S1.** Estimated values of natural history CDR-SB trajectory from 0 to 18.5 years since Aβ PET positivity.

| **Years since Aβ**<br>**PET positivity** | **CDR-SB** | **Years since Aβ**<br>**PET positivity** | **CDR-SB** |
| --- | --- | --- | --- |
| 0.0 | 0.31 | 9.5 | 2.09 |
| 0.5 | 0.34 | 10.0 | 2.51 |
| 1.0 | 0.37 | 10.5 | 2.99 |
| 1.5 | 0.41 | 11.0 | 3.54 |
| 2.0 | 0.44 | 11.5 | 4.15 |
| 2.5 | 0.46 | 12.0 | 4.82 |
| 3.0 | 0.49 | 12.5 | 5.55 |
| 3.5 | 0.52 | 13.0 | 6.34 |
| 4.0 | 0.54 | 13.5 | 7.18 |
| 4.5 | 0.56 | 14.0 | 8.07 |
| 5.0 | 0.59 | 14.5 | 9.01 |
| 5.5 | 0.62 | 15.0 | 10.00 |
| 6.0 | 0.67 | 15.5 | 11.03 |
| 6.5 | 0.75 | 16.0 | 12.10 |
| 7.0 | 0.85 | 16.5 | 13.20 |
| 7.5 | 0.99 | 17.0 | 14.35 |
| 8.0 | 1.18 | 17.5 | 15.52 |
| 8.5 | 1.42 | 18.0 | 16.73 |
| 9.0 | 1.72 | 18.5 | 17.97 |

**Figure S1.**
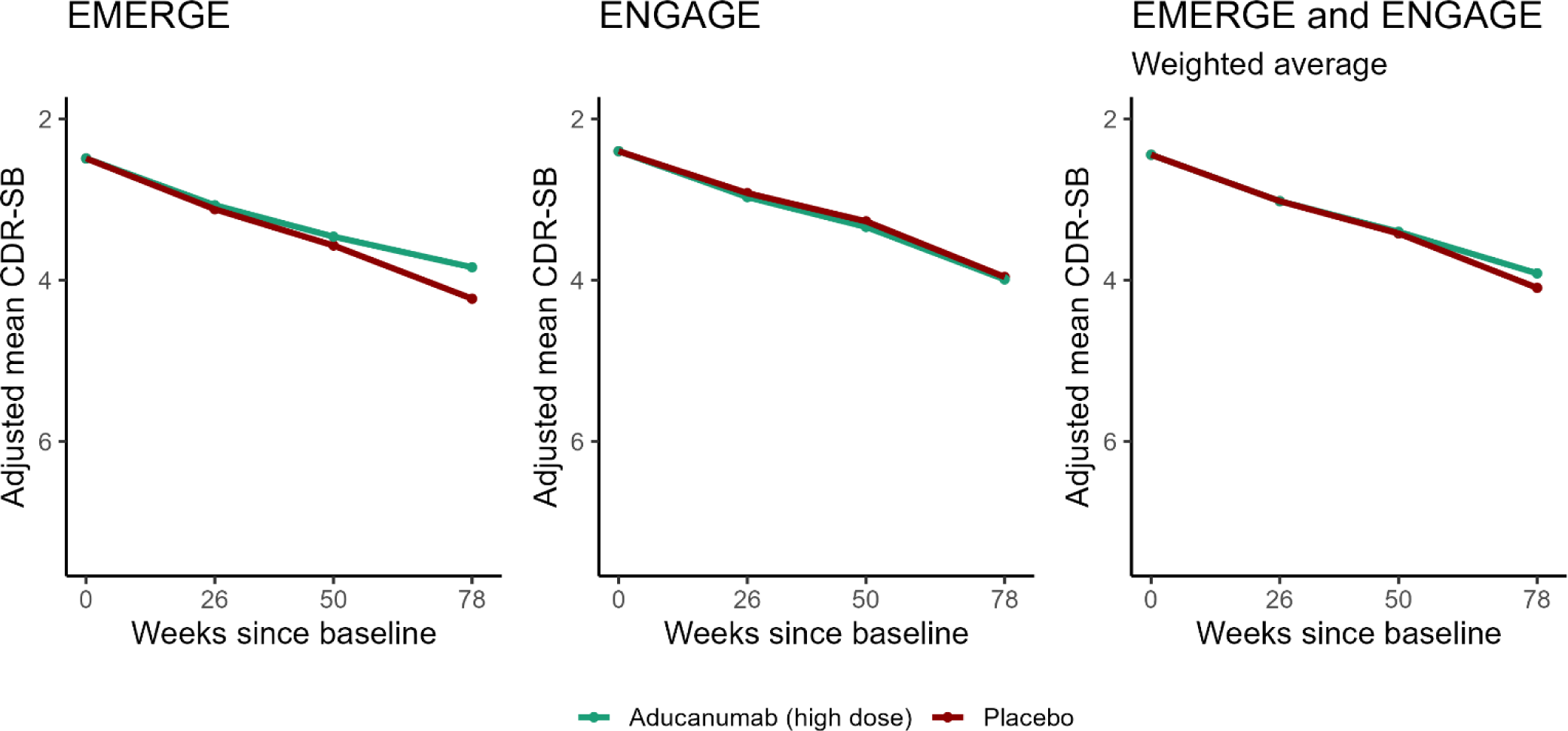
Clinical Dementia Rating sum of boxes (CDR-SB) results of the EMERGE and ENGAGE trials and a weighted average. Results are based on previously reported results of change from baseline in CDR-SB analyzed using the mixed model for repeated measures with the reported average baseline score added to the results to bring them to the CDR-SB scale. The weighted average is weighted by the baseline allocation to treatment arms.

**Figure S2.**
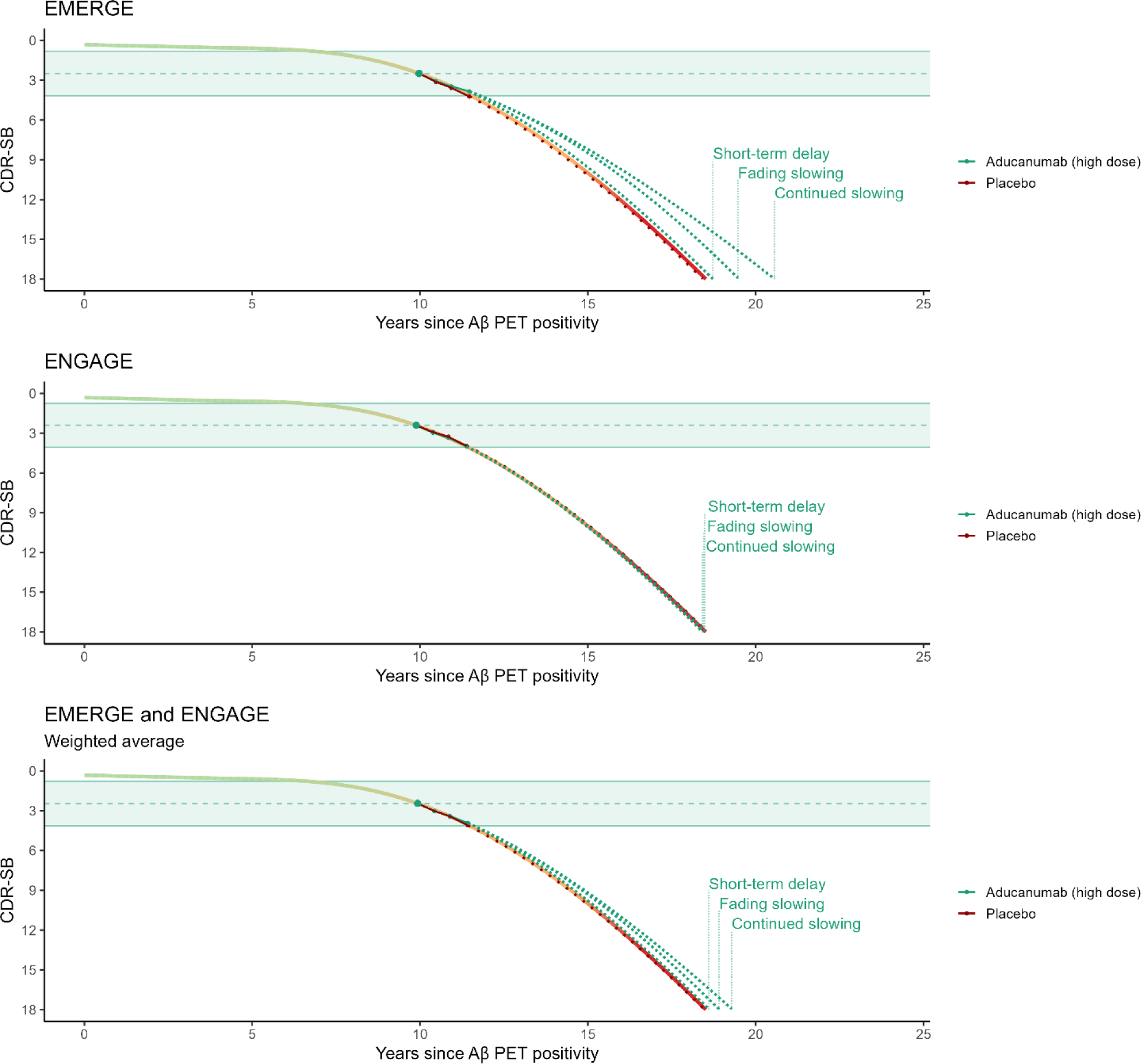
EMERGE, ENGAGE, and weighted average trial results mapped to the long-term CDR-SB trajectory with extrapolations (dotted lines). The gradient trajectory represents the estimated natural history trajectory, and the shaded bands represent the 90% prediction intervals for baseline CDR-SB scores in the different populations.

**Table S2.** Characteristics of TRAILBLAZER-ALZ 2 populations and estimated treatment effects on CDR-SB at final visit.

| **Trial** | **Treatment** | **Trial duration** | **Mean baseline CDR-SB** | **Predicted time since Aβ PET positivity at baseline** | **CDR-SB difference at final visit** | **Estimated time delay at final visit** | **Estimated time saving at final visit** |
| --- | --- | --- | --- | --- | --- | --- | --- |
| **TRAILBLAZER-ALZ 2**<br>Low/medium tau population | Donanemab | 76 weeks | 3.68 | 11.1 years | -0.67 | 30.1 weeks | 40% |
| **TRAILBLAZER-ALZ 2**<br>Low/medium/high tau population | Donanemab | 76 weeks | 3.90 | 11.3 years | -0.67 | 22.3 weeks | 29% |

**Figure S3.**
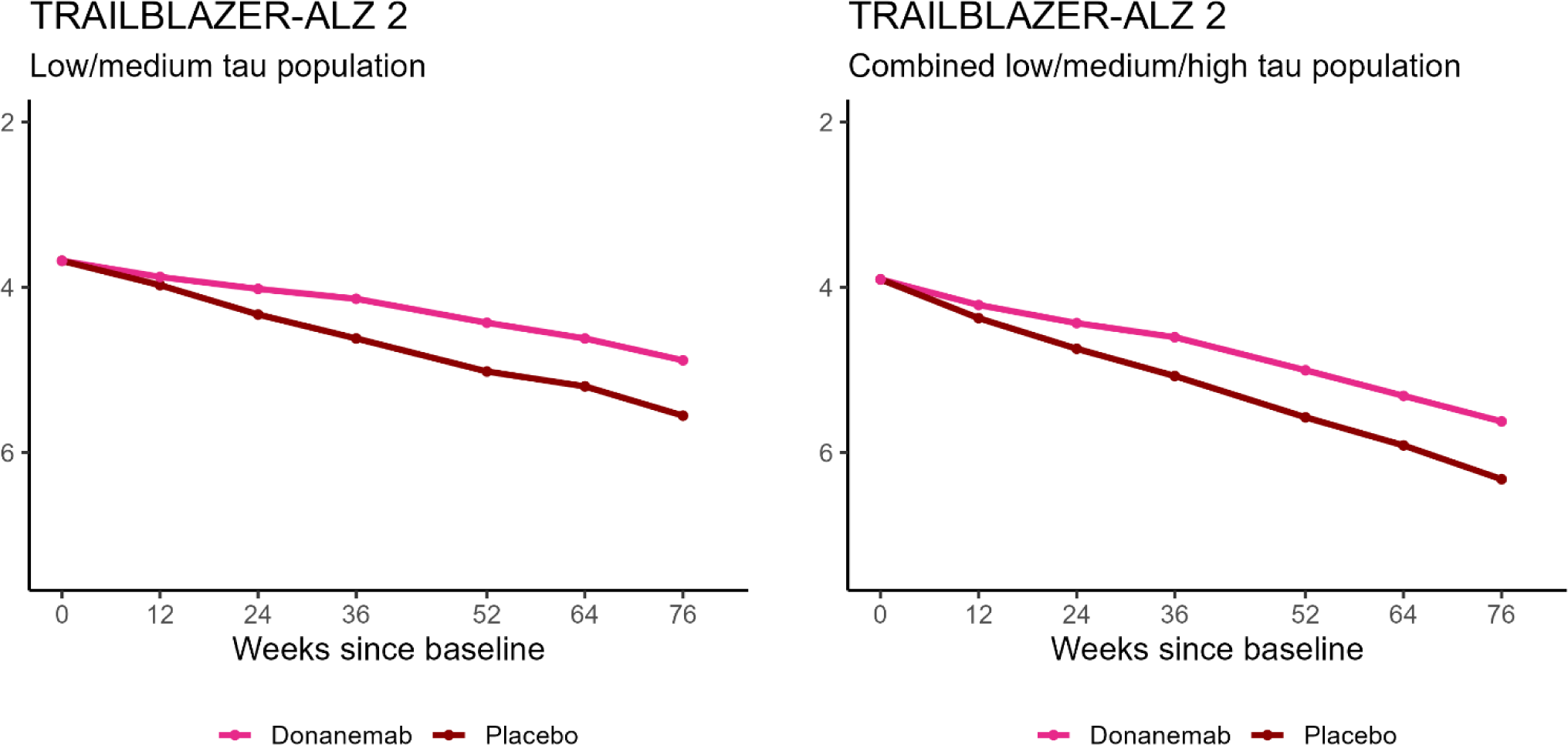
Clinical Dementia Rating sum of boxes (CDR-SB) results of the TRAILBLAZER-ALZ 2 trial for the primary low/medium tau population and the combined low/medium/high tau population. Results are based on previously reported results of change from baseline in CDR-SB analyzed using the mixed model for repeated measures with the reported average baseline score added to the results to bring them to the CDR-SB scale.

**Figure S4.**
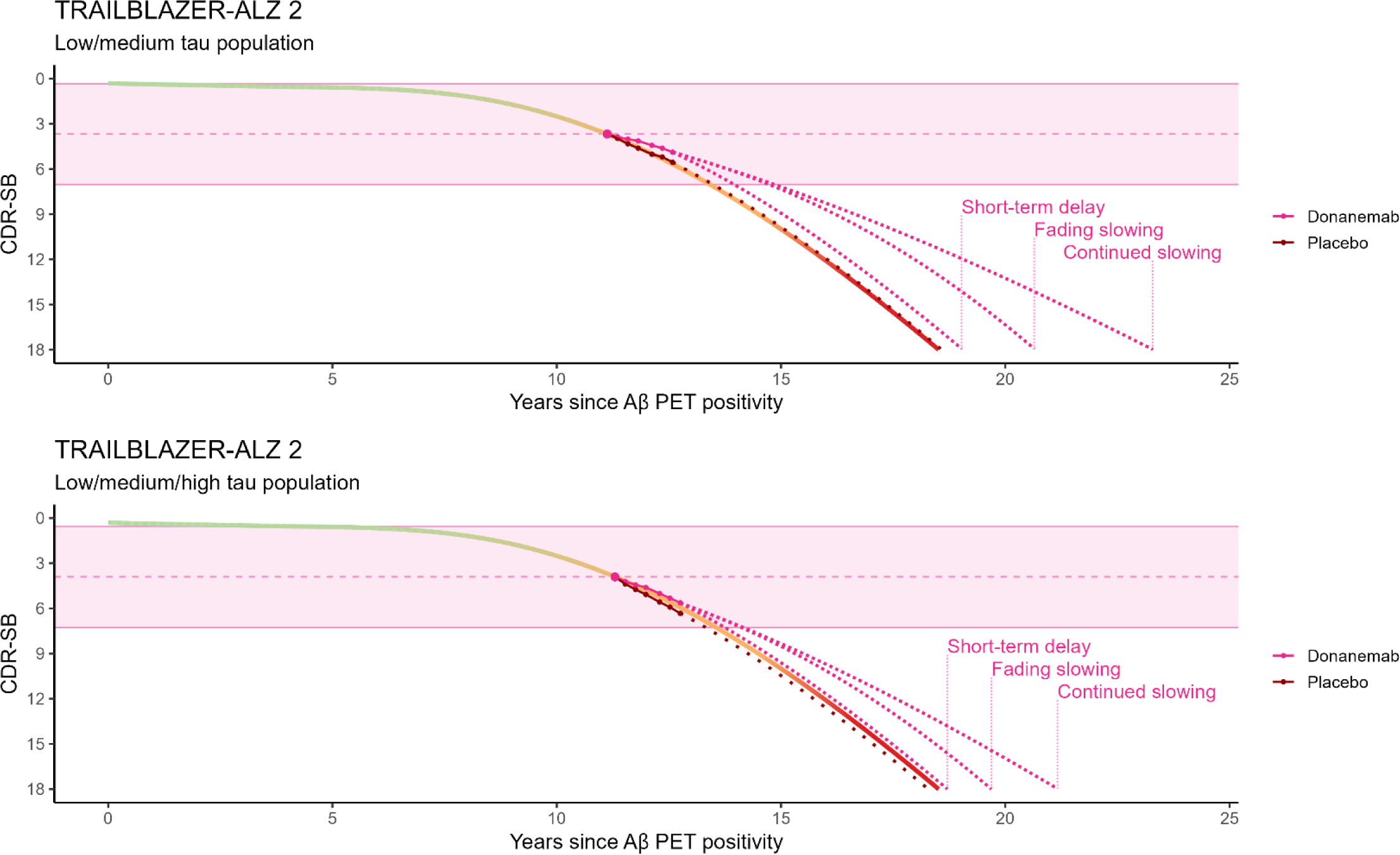
TRAILBLAZER-ALZ 2 results of the primary low/medium tau population and combined low/medium/high tau population mapped to the long-term CDR-SB trajectory with extrapolations (dotted lines). The gradient trajectory represents the estimated natural history trajectory, and the shaded bands represent the 90% prediction intervals for baseline CDR-SB scores in the different populations.

**Table S3.** Extrapolated treatment effect on CDR-SB after visit 3, 4, and 5 years for the three different scenarios across trials.

**a. Enduring short-term delay**
| Trial | Treatment | Years after initiation | CDR-SB difference to extrapolated placebo | Time delay (years) | Time saving |
| --- | --- | --- | --- | --- | --- |
| EMERGE | Aducanumab | 3 | -0.48 | 0.30 | 10% |
| ENGAGE | Aducanumab | 3 | 0.07 | -0.02 | -1% |
| CLARITY AD | Lecanemab | 3 | -0.55 | 0.47 | 16% |
| GRADUATE I | Gantenerumab | 3 | -0.32 | 0.12 | 4% |
| GRADUATE II | Gantenerumab | 3 | -0.23 | 0.21 | 7% |
| TRAILBLAZER-ALZ 2 | Donanemab | 3 | -0.84 | 0.58 | 19% |
| EMERGE | Aducanumab | 4 | -0.54 | 0.30 | 8% |
| ENGAGE | Aducanumab | 4 | 0.08 | -0.02 | -1% |
| CLARITY AD | Lecanemab | 4 | -0.61 | 0.47 | 12% |
| GRADUATE I | Gantenerumab | 4 | -0.35 | 0.12 | 3% |
| GRADUATE II | Gantenerumab | 4 | -0.25 | 0.21 | 5% |
| TRAILBLAZER-ALZ 2 | Donanemab | 4 | -0.93 | 0.58 | 14% |
| EMERGE | Aducanumab | 5 | -0.60 | 0.30 | 6% |
| ENGAGE | Aducanumab | 5 | 0.09 | -0.02 | -0% |
| CLARITY AD | Lecanemab | 5 | -0.67 | 0.47 | 9% |
| GRADUATE I | Gantenerumab | 5 | -0.38 | 0.12 | 2% |
| GRADUATE II | Gantenerumab | 5 | -0.28 | 0.21 | 4% |
| TRAILBLAZER-ALZ 2 | Donanemab | 5 | -1.01 | 0.58 | 12% |

**b. Fading stage-dependent slowing**
| Trial | Treatment | Years after initiation | CDR-SB difference to extrapolated placebo | Time delay (years) | Time saving |
| --- | --- | --- | --- | --- | --- |
| EMERGE | Aducanumab | 3 | -0.91 | 0.57 | 19% |
| ENGAGE | Aducanumab | 3 | 0.10 | -0.04 | -1% |
| CLARITY AD | Lecanemab | 3 | -1.25 | 0.87 | 29% |
| GRADUATE I | Gantenerumab | 3 | -0.39 | 0.15 | 5% |
| GRADUATE II | Gantenerumab | 3 | -0.36 | 0.28 | 9% |
| TRAILBLAZER-ALZ 2 | Donanemab | 3 | -1.83 | 1.16 | 37% |
| EMERGE | Aducanumab | 4 | -1.30 | 0.72 | 18% |
| ENGAGE | Aducanumab | 4 | 0.13 | -0.05 | -1% |
| CLARITY AD | Lecanemab | 4 | -1.84 | 1.09 | 27% |
| GRADUATE I | Gantenerumab | 4 | -0.50 | 0.19 | 5% |
| GRADUATE II | Gantenerumab | 4 | -0.54 | 0.35 | 9% |
| TRAILBLAZER-ALZ 2 | Donanemab | 4 | -2.60 | 1.55 | 35% |
| EMERGE | Aducanumab | 5 | -1.67 | 0.84 | 17% |
| ENGAGE | Aducanumab | 5 | 0.16 | -0.06 | -1% |
| CLARITY AD | Lecanemab | 5 | -2.40 | 1.27 | 25% |
| GRADUATE I | Gantenerumab | 5 | -0.60 | 0.22 | 4% |
| GRADUATE II | Gantenerumab | 5 | -0.70 | 0.40 | 8% |
| TRAILBLAZER-ALZ 2 | Donanemab | 5 | -3.33 | 1.93 | 33% |

| **Trial** | **Treatment** | **Years**<br>**after**<br>**initiation** | **CDR-SB**<br>**difference to**<br>**extrapolated**<br>**placebo** | **Time delay**<br>**(years)** | **Time saving** |
| --- | --- | --- | --- | --- | --- |
| **EMERGE** | Aducanumab | 3 | -0.95 | 0.61 | 20% |
| **ENGAGE** | Aducanumab | 3 | 0.10 | -0.04 | -1% |
| **CLARITY AD** | Lecanemab | 3 | -1.28 | 0.92 | 31% |
| **GRADUATE I** | Gantenerumab | 3 | -0.40 | 0.16 | 5% |
| **GRADUATE II** | Gantenerumab | 3 | -0.36 | 0.29 | 10% |
| **TRAILBLAZER-ALZ 2** | Donanemab | 3 | -1.88 | 1.19 | 40% |
| **EMERGE** | Aducanumab | 4 | -1.42 | 0.81 | 20% |
| **ENGAGE** | Aducanumab | 4 | 0.14 | -0.06 | -1% |
| **CLARITY AD** | Lecanemab | 4 | -1.98 | 1.23 | 31% |
| **GRADUATE I** | Gantenerumab | 4 | -0.54 | 0.21 | 5% |
| **GRADUATE II** | Gantenerumab | 4 | -0.59 | 0.38 | 10% |
| **TRAILBLAZER-ALZ 2** | Donanemab | 4 | -2.79 | 1.58 | 40% |
| **EMERGE** | Aducanumab | 5 | -1.94 | 1.01 | 20% |
| **ENGAGE** | Aducanumab | 5 | 0.18 | -0.07 | -1% |
| **CLARITY AD** | Lecanemab | 5 | -2.76 | 1.54 | 31% |
| **GRADUATE I** | Gantenerumab | 5 | -0.70 | 0.26 | 5% |
| **GRADUATE II** | Gantenerumab | 5 | -0.84 | 0.48 | 10% |
| **TRAILBLAZER-ALZ 2** | Donanemab | 5 | -3.81 | 1.98 | 40% |

## Notes

### Competing Interest Statement

LLR is an employee and shareholder of Eli Lilly and Company.
JC has provided consultation to Acadia, Actinogen, Acumen, AlphaCognition, Aprinoia, AriBio, Artery, Biogen, BioVie, Cassava, Cerecin, Diadem, EIP Pharma, Eisai, GemVax, Genentech, GAP Innovations, Janssen, Jocasta, Karuna, Lilly, Lundbeck, LSP, Merck, NervGen, Novo Nordisk, Oligomerix, Optoceutics, Ono, Otsuka, PRODEO, Prothena, ReMYND, Roche, Sage Therapeutics, Signant Health, Simcere, Suven, SynapseBio, TrueBinding, Vaxxinity, and Wren pharmaceutical, assessment, and investment companies.
AM reports no conflicts of interest.
NV has received research support from Fondation Bettencourt-Schueller, Fondation Servier, Union Nationale pour les Intárêts de la Mádecine (UNIM), Fondation Claude Pompidou, Fondation Alzheimer and Fondation pour la Recherche sur l'Alzheimer; travel grant from the Movement Disorders Society, Merz-Pharma, UCB Pharma, and GE Healthcare SAS; is an unpaid local principal investigator or sub-investigator in NCT04241068 and NCT05310071 (aducanumab, Biogen), NCT05399888 (BIIB080, Biogen), NCT03352557 (gosuranemab, Biogen), NCT05463731 (remternetug, Eli-Lilly), NCT04592341 (gantenerumab, Roche), NCT03887455 (lecanemab, Eisai), NCT03828747 and NCT03289143 (semorinemab, Roche), NCT04619420 (JNJ-63733657, Janssen - Johnson & Johnson), NCT04374136 (AL001, Alector), NCT04592874 (AL002, Alector), NCT04867616 (bepranemab, UCB Pharma), NCT04777396 and NCT04777409 (semaglutide, Novo Nordisk), NCT05469360 (NIO752, Novartis), is an unpaid national coordinator for NCT05564169 (masitinib, ABScience), NCT (AD04, ADvantage Therapeutics GmbH); has given unpaid lectures in symposia organized by Eisai and the Servier Foundation; and has been an unpaid expert for Janssen and Johnson & Johnson, all outside of this work.
MS has served on advisory boards for and received funding from Roche Diagnostics and Novo Nordisk (outside scope of submitted work)

### Funding Statement

This study did not receive any funding

### Author Declarations

This study included only published data from the following recent phase 3 trials of high-clearance Aβ-targeting antibodies: EMERGE and ENGAGE of aducanumab, CLARITY AD of lecanemab, GRADUATE I and II of gantenerumab, and TRAILBLAZER-ALZ 2 of donanemab.

